# Targeting Maternal Gut Microbiome to Improve Mental Health Outcomes-A Pilot Feasibility Trial

**DOI:** 10.1101/2024.03.08.24303670

**Authors:** Faith Gallant, Kieran Cooley, Sophie Grigoriadis, Neda Ebrahimi

**Affiliations:** NCCO Rehabilitation; CCNM; Sunnybrook Health Sciences; University of Toronto

**Keywords:** Mental health, Maternal health, Nutrition, Omega-3 fatty acids, Probiotics, Fiber

## Abstract

**Background:** Perinatal Depression and anxiety (PDA) is prevalent in new and expectant mothers affecting millions of women worldwide. Those with a history of mood and anxiety disorders are at the greatest risk of experiencing PDA at a subsequent pregnancy. Current safety concerns with pharmacological treatments have led to a greater need for adjunctive treatment options for PDA. Changes in the composition of the microbiome have been associated with various diseases during pregnancy and these changes are thought to be at least partially at play in perinatal mood disorders. While the relation between PDA and the microbiome has not been explored, evidence suggests that nutritional interventions, with fiber, fish oils, and probiotics, may play a favorable role in neuropsychiatric outcomes during and after pregnancy. The primary objective of the present study is to assess the feasibility and acceptability of a combination of non-pharmacological interventions in currently stable and pregnant women with a history of anxiety and/or depression. This study will also aim to understand ease of recruitment, treatment compliance, and protocol adherence in this cohort.

**Methods:** This a single centered, partially randomized-placebo controlled-double blind feasibility trial. 100 pregnant women, with a history of depression and/or anxiety/PDA will be recruited and randomized into one of four arms which could include: receiving a daily dose of both investigational products and dietary counselling on increasing dietary fiber, receiving a daily dose of both investigational drugs only, receiving fish oil investigational product and placebo, and a control arm with no intervention. The study involves six study visits, all of which can be conducted virtually every 3 months from the time of enrollment. At all study visits, information on diet, mental health, physical activity, and sleep quality will be collected. Additionally, all participants will provide a stool sample at each visit.

**Discussion:** It is anticipated that pregnant women with a history of depression and anxiety will be particularly interested in partaking in this trial, resulting in favourable recruitment rates. Given the positive findings of O3FA and probiotic supplements on mental health symptoms in non-pregnant adults, we expect a similar trend in PDA symptoms, with a low likelihood of adverse events. This study will build the foundation for larger powered studied to further contribute evidence for the efficacy of this potential treatment option.

**Trial Registration:** This trial was registered at ClinicalTrials/gov on October 6, 2023; NCT06074250. Trial Sponsor: The Canadian College of Naturopathic Medicine, 1255 Sheppard Ave E, Toronto, ON M2K 1E2, 416-498-1255.

## Background

Perinatal Depression and anxiety (PDA) is prevalent in new and expectant mothers affecting millions of women worldwide. Recent evidence suggests that one in seven women in the 20^th^-28^th^ week of gestation develop depression which impacts their ability to return to normal function; in the same period of gestation, anxiety has a prevalence of about 8.5-10.5% (1). Well-known risk factors include poor social support, chronic and persistent health issues in their infant, abusive partner, marital difficulties, familial history, history of violence, and other negative life events (2). The strongest predictor of perinatal depression, however, is a history of mood or anxiety disorders (2). Those with a history of depression have a 20-fold higher risk at a subsequent pregnancy (3) and relapse rates of 40% have been observed (2). Given that anxiety and depression are almost never mutually exclusive, the same risk factors likely apply to onset of perinatal anxiety. Available treatment options for these mood disorders in the peripartum population include pharmacotherapy and psychotherapy. However, safety concerns for pharmacological treatments during pregnancy and lactation, limited access to care, limited efficacy, and stigma of mental illness are major barriers to implementing these treatment options (4). Lack of treatment is associated with morbidity for the mother, infant, and family system, quality of life impairment, and inadequate response to presently available treatment options (4). Thus, a need for adjunctive treatment remains, for optimal management of perinatal anxiety and depression disorders.

The Gut Brain Axis (GBA) is a recently realized phenomena in medicine, suggesting continuous bidirectional communication between the gut microbial community and the brain; a requirement in achieving and maintaining gastrointestinal (GI) homeostasis and cognitive function. Dysbiosis-an imbalance in bacterial composition-has been implicated in many disorders, including mental health (5).

Pregnancy itself has been associated with changes in gut microbiome driven by immunological, hormonal, and metabolic changes-these changes are pro-inflammatory in nature, leading to increase in cytokines and leukocytes and recruitment of other immune cells (macrophages, Natural killer Cells, etc.) that aid in angiogenesis, transport of respiratory gases and nutrients between mother and fetus, and protect against pathogens (6). Interestingly certain changes in the composition of the microbiome have been associated with various diseases during pregnancy such as gestational diabetes, preeclampsia, fetal growth restriction, and obesity (6). Likewise, these changes are thought to be at least partially at play in perinatal mood disorders. Despite documented pregnancy specific alterations in maternal microbiome (6), the relation between PDA and microbiome has not been explored.

A 2022 mini review examined the correlation between perinatal depression and dysbiosis of the mother’s microbiome and reported dysbiosis as a likely precipitating factor in the development of psychiatric disorders during pregnancy (7). Emerging evidence suggests that nutritional interventions, with fish oils, probiotics and prebiotics, may play a favorable role in neuropsychiatric outcomes (8-13) and that some of these effects are mediated at least partially, by restoring eubiosis.

The protective effects of specific nutrients and diets have been demonstrated in major depressive disorder (MDD). For example, the onset of depression has been shown to increase with high fat western diets and significantly reduce with Mediterranean diets (5). The latter has demonstrated a reduction in oxidative stress, and increase in neurotransmitters serotonin, noradrenalin, dopamine, and monoamines, all of which play a role in major depression (5). The higher intake of plant-based fiber and omega-3 fatty acids (O3FAs) in Mediterranean diets may partly be responsible for this protective effect. Dietary fiber is fermented by cecum and large intestine bacteria. In addition to changing the abundance of specific strains of bacteria, it also drives the levels of short chain fatty acid (SCFA) production (14). Several observational studies have linked fiber intake to reduction in severity of depression as well as depression onset. Mechanisms proposed for this outcome include increases in levels of certain neurotransmitters, Brain Derived Neurotrophic Factors (BDNFs) and reduction in inflammatory biomarkers (14).

Specifically in the pregnant population, a recent retrospective cohort analysis examined the results of three studies (a randomized controlled trial of a low-glycemic-index diet in pregnancy, the pregnancy exercises and nutrition research study, and a randomized controlled trial on probiotics). Together, data from 1521 participates were included in this analysis which showed that the average daily intake of fiber was statistically significant with regard to maternal well-being (r=0.13; P<0.01). Results noted that nutrients in whole grains, fruits, and vegetables were associated with improved mental health, and the authors suggested that the high fiber content in such foods is the sources of these observed benefits (15).

Multiple randomized controlled trials (RCTs) have investigated probiotics in pregnancy. In 2017, Slykerman et al., reported significantly lower postpartum depression and anxiety scores in women taking probiotics versus placebo (16). However, this analysis was done retrospectively as a secondary outcome. A 2020 RCT by Dawe et al., found no impact in psychiatric outcomes of obese women at 26 weeks of gestation. However, this sample had lower baseline mental illness scores than what would have been expected, and there was no postpartum assessment (17). In 2021, Browne et al., completed a pilot RCT and concluding that the impact of probiotics on prenatal maternal anxiety and depression is feasible and acceptable. While depression and anxiety symptoms decreased after intervention, authors found no significant difference compared to placebo, and opined that the small sample size (n=40) may have underpowered the design (18).

Recent evidence points to O3FAs having an impact on the composition of the microbiome. In rodent models, maternally-separated rats with increased stress markers, have an altered composition of bacteria in their microbiome, including decreased numbers of bacteria in the *Lactobacillus* genus and elevated numbers of the *Oscillibacter, Anaerotruncus* and *Peptococcus* general (19). Pusceddu et al., showed that with long-term administration of eicosapentaenoic acid (EPA)/docosahexaenoic acid (DHA) in stressed rodents, there is a restoration of the microbiome composition and reduction of inflammatory processes typically associated with stress (20). Specifically, EPA/DHA supplementation appeared to restore the microbiome composition back to a state similar in non-stressed rodents including increased *Lactobacillus* genus and reduced *Anaerotruncus* genus (20).

Currently data linking O3FA administration with benefits in mood disorders through microbiota modulation is mostly limited to rodent models. However, the positive findings from observational and clinical trials on the impact of O3FAs on mood disorder, call for a closer look at the interaction of fish oil with microbiome and impact on the GBA of pregnant women (21).

### Rationale for study

Evidence for the positive role of fiber, O3FAs and probiotics in anxiety and depression is accumulating. Studies on the combination of these treatments, and their mitigating effects on the microbiome remain elusive. Furthermore, a significant knowledge gap remains, on their usability and efficacy in the pregnancy population, particularly in relation to mental health disorders. Given the non-pharmacological nature of these interventions, their wide availability, safety, and cost effectiveness, they have a high potential to serve as an effective and preferred adjunctive treatment for management and/or prevention of mental health disease in pregnant and lactating mothers.

#### Objectives

The main objective of this study is to assess the feasibility and acceptability of a combination of non-pharmacological interventions - high fiber diet, probiotics, and fish oil supplementation-in currently stable and pregnant women with a history of anxiety and/or depression. This population has specifically been chosen as a history of mood or anxiety disorders is the strongest predictor of PDA (2).

This study will also aim to understand ease of recruitment, treatment compliance, and protocol adherence in this cohort. The findings of these objectives will inform on strategies needed to build a larger trial, aimed at comparing efficacy of these interventions and the magnitude of change and impact of the microbiome and PDA.

## Methods

### Trial Design and Setting

This a single centered, partially randomized-placebo controlled-double blind feasibility trial with three intervention arms (Gutopia, Gutboost, and Gutless) and one active control. 100 pregnant women, with a history of depression and/or anxiety/ PDA attending Sunnybrook Hospital Clinics, in the Greater Toronto Area will be recruited.

Participants in Gutopia will receive a daily dose of both investigational products and dietary counselling on increasing dietary fiber. Participants in Gutboost will receive a daily dose of both investigational drugs, while participants in the Gutless arm will receive fish oil and placebo. Participants in Gutless and Gutboost will be blinded to the status of placebo/probiotic they will be receiving. The study member interacting with the patients will also be blinded to the treatment group the participant is assigned to. Participants who fully consent to receiving both investigational products including placebo, will be fully randomized to one of the intervention arms. Participants not willing or able to take placebo or probiotics will be assigned (not randomized) to the appropriate group that accommodates their preference/needs. Participants unable to or unwilling to take any of our investigational products will be assigned to the active control arm. Women in the control arm will receive standard care, offered to all obstetric patients seen at the hospital.

The study involves six study visits, all of which can be conducted virtually every 3 months from the time of enrollment (Figure 1). The last study visit will happen between 9-12 months postpartum. Two of the study visits will occur during pregnancy (Enrollment (Visit 1) and 3^rd^ trimester (Visit 2)) with the remaining occurring at 1, 3, 6 and 9 months postpartum. At all study visits, information on diet, mental health, physical activity, and sleep quality will be collected. Additionally, all participants will provide a stool sample at each visit, using our stool collection kits, which will be shipped to, or dropped off at our center.

**Fig. 1.**
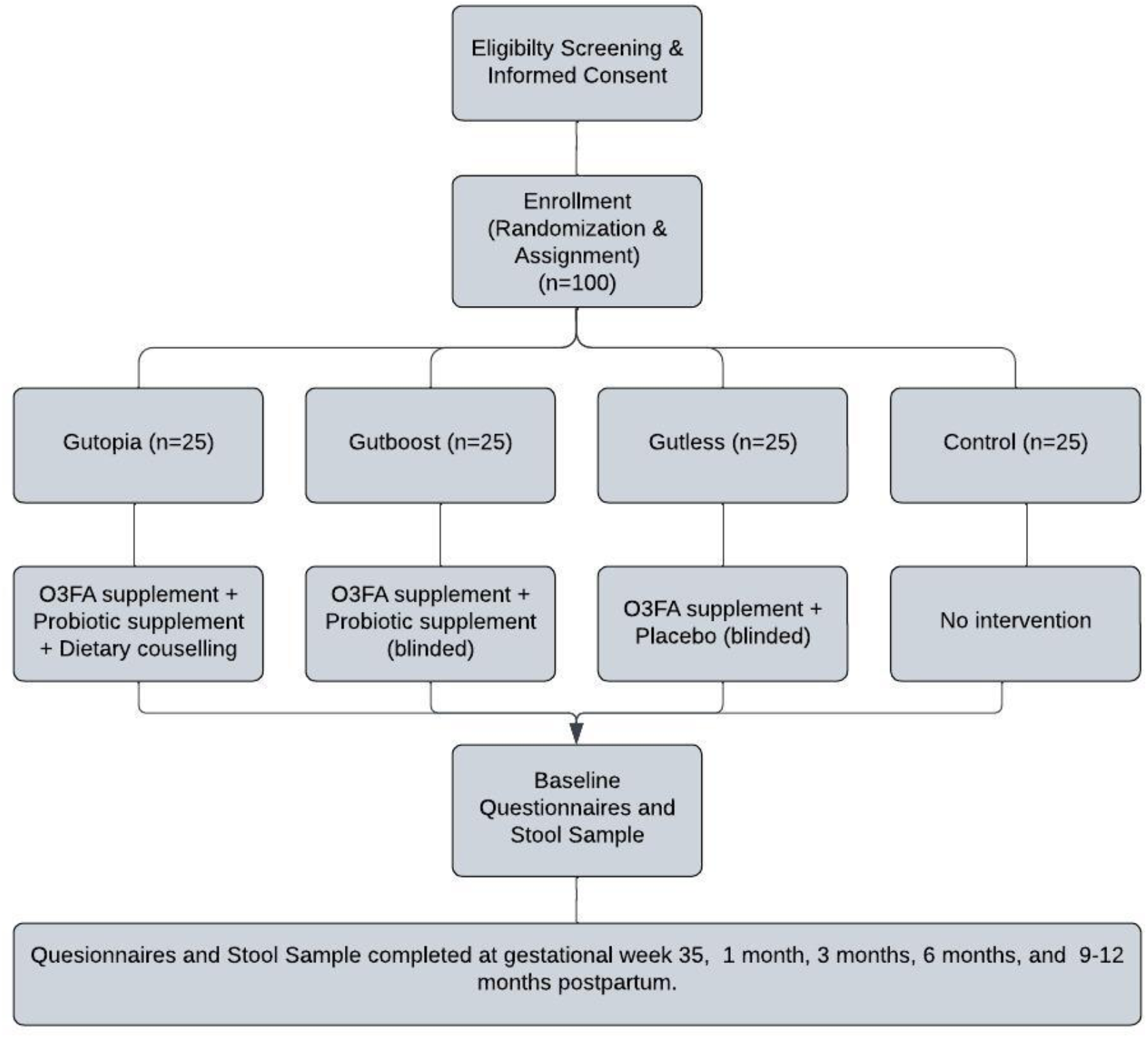
Diagram of participant flow through the trail.

The study has been reviewed by the Research Ethics Board of the Canadian College of Naturopathic Medicine and the Research Ethics Board of Sunnybrook Health Sciences Centre. Any amendments to this protocol will be communicated to the Research Ethics Board and Health Canada (the authority responsible for approval, conduct, oversight, and inspection of clinical trials in Canada) and modified on the trial registry.

### Participant Eligibility

#### Inclusion Criteria

1. A person aged 18-43 years old.
2. At 12-35 weeks of gestation at time of enrolment.
3. Uniparous pregnancy.
4. A non-smoker, alcohol, or recreational drug user.
5. Financially stable.
6. In a married or common law relationship
7. Clinical diagnosis of lifetime or current depression/anxiety or PDA, but stable at the time of recruitment.
8. No other significant comorbidities.
9. Ability to read in English and provide informed consent.

#### Exclusion Criteria

1. Pre-pregnancy BMI>30.
2. Low income (unable to afford basic daily needs)
3. Single parent without any kind of family support
4. Having a child with significant mental/physical disability.
5. Diagnosed with other major mental health disorders.
6. Taking prescription medications (other than SSRIs/SNRIs/TCA).
7. Smoking and recreational drug use
8. Dietary restrictions or allergies to fish oils.
9. Repeated antibiotic use.
10. Women unable to switch to study brand supplements.
11. Multiparous women with young children (i.e., one or more children less than 4 years of age at the time of delivery).
12. No/low English reading comprehension

Our inclusion criteria allow for antidepressant/anxiety use. Medication use is an important prognostic factor that can significantly alter mental health outcomes. Hence the number of patients treated with an antidepressant/anxiolytic needs to be equally distributed between all arms. To achieve this, we will use stratified randomization using two strata (treated vs. untreated) through a computer-generated process using permuted blocks of three and six. The process will be repeated separately for untreated subjects.

### Sample Size, Recruitment Plan, & Pre-study Screening

While a consensus on an ‘ideal’ sample size for pilot/feasibility studies hasn’t been reached, our proposed number meets recommendation for best practice for conduct of pilot trials.

Pregnant patients attending The Women’s Mood and Anxiety Clinic at Sunnybrook Hospital, will be informed by their psychiatrist about the study and permission to share their contact number with the research team will be requested. Interested patients will be called by a member of the research team and screened for inclusion/exclusion criteria and their ability/status on O3FA and probiotic intake. Patients not willing or unable to take either supplement, will receive information about the control arm of the study. Patients willing to take the study supplements, will receive information about the interventional arms of the study and the process of randomization. Subjects will also be informed that they may receive placebo instead of probiotics. This design accommodates patient preferences and expectations and emphasizes the goal of an inclusive, pragmatic study design to generate real-world data.

### Randomization & Baseline Evaluation

All eligible patients interested to partake, will be consented, and randomized or assigned to an intervention or control arm based on responses during screening. If eligible for full randomization, this will be completed by the Study Principal Investigator, using a computer-generated randomization via random permuted blocks. The study coordinator will only be informed of the status of randomization to the Gutopia arm and will be blinded to the Gutboost and Gutless arms.

Once randomized, the participant will be scheduled for the baseline assessment as soon as possible. If scheduled as a virtual visit, a stool kit will be mailed to the participant or provided at their prenatal visit. There is no predetermined time between the screening and baseline, however the baseline visit will take place prior to the 35^th^ gestational week (visit 2). Collection of baseline questionnaires will include the following: The Generalized Anxiety Disorder-7 (GAD-7), The Edinburgh Postnatal Depression Scale (EPDS), Single Item Sleep Quality Scale (SISQS), Pregnancy Physical Activity Questionnaire (PPAQ), Consumer Financial Protection Bureau - Financial Wellbeing (CFPB-FWB), Demographic Questionnaire, and the Dietary Screening Form. The baseline visit will also include provision of a stool sample.

There will be at total of six study visits with the remaining four taking place in the postpartum period (Figure 1). All questionnaires completed at baseline, with the exception of the CFBP-FWB, the Demographic Questionnaire and the Dietary Screening Form, will be completed at all postpartum visits. A Post Delivery Questionnaire will be completed at visit 3 (∼4-6 weeks postpartum), and postpartum questionnaire will be completed at visits 4-6. The sessions will be 45-60 minutes in length.

### The Intervention

Participants in all intervention groups will receive a 200-milliliter bottle of Genestra brand Super EFA Forte Liquid Omega-3 Fatty Acid. Participants will be instructed to take 1 teaspoon a day with a meal.

Participants in the Gutopia and Gutboost intervention groups will receive HMF Maternity Probiotic Formula, a Genestra Brand. Participants will be asked to take one tablet daily with food.

Details of both supplements are provided under the ‘Investigational Products’ in the following section.

Only participants in Gutopia will receive dietary intervention. Intervention is geared to increase gut friendly foods via a flexible dietary plan aimed to increase daily uptake of prebiotic and probiotic foods. A dietician will use the baseline food questionnaire data provided by each subject at enrollment to devise strategies and recommendations for increasing intake of pre/probiotic foods. The target goal is to include at least 35 grams/day of fiber and at least 1-2 servings of fermented foods daily. The dietary intervention will also include recommendations minimizing highly processed foods, high in sugar and fats. To improve adherence and accountability, weekly follow-ups by phone, virtual meeting or email will be conducted by a trained study team member until patients have reached a level of confidence in attaining goals. During each follow-up the study team will monitor level of adherence to the study dietician’s recommendations. These follow-ups will be designed to provide encouragement while helping patients address and prevent triggers that jeopardizes their fiber intake.

#### Use of Theory

Given the nature of this feasibility study, we aim to promote increased accountability with regular follow-ups to drive change. To achieve this, theoretical proponents of motivational interviewing (MI), will be incorporated into study visits. For instance, behaviour change will be enabled by causing participants to verbalize arguments for changes, and decrease language which favors the status quo. Further, relational factors, such as expressions of empathy will be used to promote positive change (22).

#### Educational material and resources

Participants in the Gutopia arm will be presented with food tables listing common foods, and snacks from different food groups, containing the highest amount of fiber per serving. This is to help enable participants reach their target daily fibre with the fewest servings possible. These tables are categorized by food group, and providing the food name, brands and recommended serving sizes. These have been provided in the supplemental information section.

#### Investigational Products

All subjects in treatment groups will receive a 200-milliliter bottle of Genestra brand Super EFA Forte Liquid Omega-3 Fatty Acid. This product contains a total of 2600mg of O3FA (1500mg of EPA + 1000mg of DHA) per 5ml serving. (Participant will be instructed to take 1 teaspoon a day with a meal. Each bottle contains about 40 servings and with a daily consumption of 1 teaspoon a day, expected to last 6 weeks. Each patient will receive supplies for the duration of the study period. While no clear upper tolerable dose in pregnancy has been identified, no increase in risk of bleeding or complications at delivery have been observed in pregnant women (n=533) receiving 2.7g/day during the last trimester compared to women receiving olive oil or no supplements (23,24). The dose used in our trial is consistent with a dose that is likely effective and is below the safe maximum dose suggested in these trials.

We will use HMF Maternity Probiotic Formula, a Genestra Brand. Each capsule contains 10 billion of four strains of bacteria: *Lactobacillus Salivarius* (6.25 billion CFU), *Bifidobacterium animalis subsp*.*lactis & Bifidobacterium bifidum* (2.5 billion CFU) and *Lactobacillus Paracasei* (1.25 billion CFU). Meta-analysis of several RCTs of probiotic use in pregnancy reported no increased risk to fetus (25). Furthermore, the probiotic supplement in our trial was specifically labeled for pregnant women, following its use in a double-blind, placebo-controlled clinical trial of 454 pregnant women-no increase in adverse events in comparison to placebo was noted (26).

The placebo capsules will contain [48% Maltodextrin, 48% Microcrystalline cellulose, 2% Magnesium stearate, 2% Silicon dioxide]. The Capsule shell will be composed of Hydroxypropyl ethylcellulose and will be identical in capsule shape and packaging to the probiotics. Subjects will be instructed to take one capsule a day with food, 2-3 hours before or after taking an antibiotic. Each box of probiotics/placebo is expected to last 30 days. We will provide enough supplies for the duration of study period.

#### Concomitant Therapies

All concomitant medication, psychotherapy, or natural health products will be allowed during this trial. Participants will be encouraged to continue all other previous treatments at the same dose for the duration of the trial, if possible, but allowed to make changes if recommended by their mental health care provider. Participants will be queried at baseline and each follow-up visit about their use of medication, psychotherapy, or natural health products, and any changes will be recorded, analyzed, and reported.

#### Safety profile of supplements

Marangell et al., studied the impact of O3FA supplementation (2960 mg/d) on the prevention of postpartum depression. Adverse events reported were mild. 4 participants reported a “fishy” after taste, 1 reported mild dyspepsia and 1 reported increased stool frequency with loose stools. There were no withdrawals due to adverse events (27). Similarly, Su et al., completed an RCT with 24 participants. No withdrawal of participants due to adverse events was reported. 12/18 participants in the placebo group and 10/18 in the O3FA group did not report any adverse events. Events reported included insomnia (2 in the placebo group and 3 in the O3FA group), nausea (4 in the placebo group and 6 in the O3FA group), and diarrhea (2 in the placebo group and 1 in the O3FA group). The authors noted that most events were mild with the exception of significant nausea for 1 participant in the placebo group which lead to termination of treatment. The authors reported no effects on biological parameters including abnormal bleeding time or liver function. All newborns were in normal general health at birth (28). Further Mattes et al., studied O3FA supplementation (3.7 g/d) in 83 pregnant women in the prevention of maternal depression. The authors documented no adverse events (29).

2020 meta-analyses by Zhang et al, examined the efficacy and safety of EPA and DHA monotherapy in the treatment of perinatal depression in women with a clinical diagnosis of MDD or symptomatic depression in pregnancy and/or postpartum. Only randomized double/triple blinded placebo-controlled trials were included in this study (N=8) (30). A total of 638 pregnant women were administered 1-6g/day a day. The safety of doses of 1-6 grams/day were demonstrated in these trials and did not differ significantly between O3FAs and placebo groups within each trial. The most reported side effects were mild and self-limiting. They included: nausea, vomiting, increased stool frequency, dizziness, fatigue, and insomnia.

The safety of probiotics has been established. The theoretical risks include: infection, adverse metabolic activities, excessive immune stimulation and gene transfer, in susceptible immunocompromised individuals. As far as strains specific to the investigational products: no safety concerns nor adverse effects were observed in clinical trials administering Lactobacillus salivarius CECT5713 (31).

No safety concerns have been reported in studies administering Lactobacillus Paracasei LC01 (32), Bifidobacterium animalis subsp. Lactis (33) & Bifidobacterium bifidum (34) to healthy adults.

Long-term safety and efficacy of probiotics administered during the perinatal period was established in a randomized double-blind placebo-controlled study of over 300 pregnant women, involving strains: *Lactobacillus rhamnosus GG, Bifidobacterium lactis Bb-12, Lactobacillus paracasei ST11, and Bifidobacterium longum BL999* (35). Authors found no differences in growth or non-communicable disease prevalence between children receiving perinatally probiotics or placebo.

The safety profile of probiotics is deemed excellent in healthy adults and pregnancy is not considered a contraindication for their use.

#### Criteria for Study Withdrawal

All eligible participants will be informed that their participation is completely voluntary, and they can discontinue participation at any time. Subjects may withdraw from study entirely or from specific parts of the study only. If the latter is desired, then follow-up pertaining to other consented components of study will continue.

Data collected prior to withdrawal may be retained and used in a manner consistent with study purpose and protocol. The exception is if subjects explicitly request the removal of all previously collected information and data, in which case the study team will oblige. Once a participant expresses a desire to withdraw from the study, the research team member in contact with the participant will confirm whether previously collected data can be retained (or should be destroyed), and whether the participant can be contacted in the future for other research opportunities.

The participant will be notified during consent and at the time of withdrawal that any data retained for the analysis, cannot be withdrawn after study closure or publication of study (whichever comes first). Lost to follow-up is defined as enrolled participants that were randomized and completed all parts of Visit 1, but complete zero parts of any subsequent Visits (2-6). Five attempts will be made to reschedule the study visits for these participants. The number of attempts and mode of communication to reach these participants will be documented. If the missed Study visit is not successfully re-scheduled within 4 weeks, the Visits will be marked as lost to follow-up. Should the participant return to completing the later visits, they will be included in all analysis consistent with intention to treat. However, if all attempts to reconnect by the study team fail for the missed visit and all remaining visits, and/or should the participant explain that they wish to stop participating in the study, they will be marked as Study Withdrawal.

All adverse events during the study period will be recorded, and these records will be maintained for 15 years. The clinical trial sponsor will inform the Natural and Non-prescription Health Product Directorate of Health Canada of any serious expected or unexpected adverse events related to the study product immediately if possible and no later than 7 days after becoming aware of the information. Reported symptoms of psychosis (hallucinations, delusions, or disordered thinking), will trigger a referral of participant to the study psychiatrist for assessment and whether they meet the criteria for withdrawal. If a participant reports a significant worsening in symptoms of anxiety or depression, they will be permitted to remain enrolled in the study but will be advised to contact their primary mental health care provider for assessment and management. If the mental health care provider suggests that psychiatric rescue medication is needed, this clinician will recommend the treatment to the participant. Changes to treatment regimens must be reported to study team for documentation.

Serious adverse events will also be reported to the Research Ethics Board which has reviewed the study protocol.

Participants that complete all parts of the study but fail to adhere to regular supplemental intake (miss 7 or more consecutive days), will be included in the study for as long as they wish. Their reasons for not taking the supplements will be documented and strategies to help overcome this (i.e. forgetfulness) will be made. If reasons for stopping supplements are allergic reactions or side effects, they will be withdrawn from the intervention and an adverse report will be made consistent with the safety protocol. If participants still wish to continue with the study, they may do so as part of the control arm.

#### Outcomes Measured

As a feasibility study, process evaluation will be our primary outcome. In the first 6 months of the trial, we will monitor number of patients screened, randomized, and enrolled per month, average time lapse between screening and enrollment, and time taHken to enroll 15% of our target (i.e., n=15). We will also assess baseline rates of O3FA and probiotics supplementation in this specific cohort prior to randomization. We will identify and document all barriers in meeting target numbers needed for each study arm and address them appropriately. In patients successfully enrolled, we will also monitor compliance, side effects and challenges in taking supplements in the intervention groups. We will closely document all issues in adhering to intake instructions and will attempt to resolve them on a case-by-case basis (i.e., if forgetfulness is a cause, daily automatic email reminders will be sent to that individual). In Gutopia, attaining daily fiber target of 35 grams/day will be monitored frequently via phone call in the first six months or until the patient no longer needs the support. Finally, we will assess retention rates, proportion of study visits completed and difficulty in the collection and shipment of stool samples and completion of questionnaires and dietary recall. All barriers to timely completion of these tasks will be documented and managed on a case-by-case basis. Strategies to overcome each of these barriers will be proposed in the final write-up of this study.

As our secondary aims, we will seek the opportunity to produce preliminary data on the association between maternal microbial response to each intervention, and self-reported scores on questionnaires of mental health challenges. This will be for exploratory purposes to identify any signals that will further support the conduct of a larger trial.

### Mental Health Outcome

The EPDS and GAD-7 scores will be derived for each subject at enrollment and at every subsequent follow-up. We will calculate mean and median scores for each group and conduct between group analyses. Additionally, we will compare the changes between baseline and follow-up scores for each group (within group analysis); followed by a between group comparison of these changes.

#### Microbiome Profiling

All stool samples collected at each study visit will be stored in -80 degree Celsius freezers. At the end of the study period, all samples will be shipped to Microbiome Insight, in Vancouver BC. All samples will be analyzed as one batch, using shallow shutgun metagenomic sequencing, to reveal the composition (diversity and abundance of strains at taxomonic levels) and metabolic profile (functional analysis) of the gut microbiome This procedure will be completed by a bioinformatics specialist at Microbiome Insigt-blinded to all aspect of study.

### Microbial Response to Diet and Supplementation

First, we will assess microbiome response to each intervention within each group, by comparing baseline (pre-intervention) and follow-up visits (Visit 2-6) changes in microbiome composition. The gut profile will be compared between all four arms at each follow-up. The within group changes from baseline will be compared between groups.

### Microbiome and Mental Health

We will measure the interaction between diet and supplementation. The objective is to understand the differences in microbial and metabolic profiles of each group in response to the interventions and the corresponding changes in mental health scores.

### O3FA & Probiotic Interaction

The synergism between O3FAs and probiotics has not been defined. We expect that comparing the microbiome between Gutboost and Gutless will yield differences between O3FA alone and O3FA + probiotics on mental health scores.

#### Statistical Analysis

Should the trial succeed, and we continue to recruit for a larger trial, the following statistical plan will be used to compare mental health outcomes between the groups. We will use mental health scores as primary outcome and an intent to treat approach for these analyses.

Participants lost to follow-up will be assumed to have lost interest /commitment to the study and will be excluded from the analysis. Those that discontinue treatment will still be included.

Between group differences for EPDS/GAD-7 scores will be analyzed using Analysis of Covariance (ANCOVA) with group membership as the independent variable, EPDS/GAD-7 as dependent, pharmacotherapy, exercise status, and Cognitive Behavioral Therapy (CBT) as covariates.

The interaction effect between O3FA and probiotics model will be analyzed and reported in each model. Alternative models will be used if the assumptions of ANCOVA are violated. If differences are found between groups, appropriate post-hot analyses will be conducted to identify where the differences lie. Within group differences at each time point will be analyzed using Repeated Measures ANOVA. Data from each follow-up will be compared to baseline scores. EPDS/GAD-7 scores will be the dependent variables and follow-up times will be our independent variable.

Comprehensive shotgun bioinformatics will be conducted by Microbiome Insight to provide details on taxonomic and community composition, diversity and similarity of functional profiles, analysis of variation, and differential abundance. The relationship to each intervention and relationship to EPDS and GAD-7 scores will be analyzed and reported by an experienced bioinformatician.

#### Data Management

The collection and storage of data will be done according to Good Clinical Practice Guidelines. All study staff involved in the collection or entry of data will be trained by the trial coordinator and will complete ethical conduct of research training (Tri-Council Policy Statement: Ethical Conduct of Research Involving Humans 2). Data from potential and enrolled participants will be stored in the secure platform, Research Electronic Data Capture (REDCap), and a password-protected database on a secure server managed by the Canadian College of Naturopathic Medicine. Data collected at each study visit will be captured using an electronic case report form which will be entered into the database. Questionnaire completion will be done by the study participants through REDCap. Laboratory data will be entered into the database and double-checked for accuracy by a team member blind to participant allocation. An audit of the data will be completed by a senior member of the research team at 3 months. All information obtained during the trial will be kept in a locked cabinet or on a password-protected computer and destroyed (deleted or shredded) after 15 years.

#### Access to data and dissemination

All personal health information will be kept confidential and only accessed by the research team, unless required by law. This includes an audit by the Canadian College of Naturopathic Medicine Research Ethics Board or the Sunnybrook Health Sciences Center Research Ethics Board or Health Canada Natural and Non-prescription Health Products Directorate. All data will be maintained on password-protected, secure servers or locked filing cabinets for the required length of time. Following this time, data will be destroyed securely by deleting the digital files or shredding paper documents. No identifying information from any participant will be used in the dissemination of study results. Dissemination of the study findings will occur through publication in open access journals. Study participants will have the option to be notified of the study findings.

## Discussion

Given the prevalence of PDA and its resulting impact on quality of life and infant well-being, the importance of investigating underutilized and overlooked non-pharmacological treatments is paramount. Specifically, the need to investigate adjunctive treatment approaches in comparison to prescription medication and psychotherapy has been identified as an important need by expecting and new mothers. It is anticipated that pregnant women with a history of depression and anxiety will be particularly interested in partaking in this trial, resulting in favourable recruitment rates. Given the positive findings of O3FA and probiotic supplements on mental health symptoms in non-pregnant adults, we expect a similar trend in PDA symptoms, with a low likelihood of adverse events.

While not our main objective, we anticipate a correlation between the microbial composition and O3FA and probiotic supplementation. As several recent studies have demonstrated a positive impact of probiotics or O3FAs on mental health outcomes via changes in the gut microbiome, this has yet to be demonstrated in individuals at risk of PDA. Having intervention groups which provide both supplements (Gutboost) versus O3FA alone (Gutless), will allow us to determine potential interaction between these supplements. We anticipate a difference in mental health scores between the Gutboost and Gutless intervention groups. Incorporating a third intervention arm where dietary counselling is provided in addition to supplementation (Gutopia), further allows us to understand the interaction between diet and supplementation. We anticipate differences in mental health scores between Gutopia compared to the other intervention arms. Additionally, having an active control arm, can potentially inform on the magnitude and directionality of these changes by comparing outcomes in the intervention arms (N=75) to the active control arm (N=25).

As this feasibility trial is the first to study O3FA and probiotic supplementation for PDA symptoms, it will build the foundation for larger powered studied to further contribute evidence for the efficacy of this potential treatment option. This type of evidence creates a rationale for the inclusion of nutrition professionals in mental health care teams and the use of dietary counselling in the treatment of PDA. Nutrition interventions can be low in risk, acceptable to patients, cost-effective, and may have additional benefits to overall health, which is particularly important for our target population.

## Supporting information

Randomization Flow Chart

## Data Availability

All data produced in the present study are available upon reasonable request to the authors

## Abbreviations

ANCOVA: Analysis of Covariance
BDNF: Brain Derived Neurotrophic Factors
CBT: Cognitive Behavioural Therapy
CFPB-FWB: Consumer Financial Protection Bureau - Financial Wellbeing
DHA: Docosahexaenoic Acid
EHA: Eicosapentaenoic Acid
EPDS: Edinburgh Postnatal Depression Scale
GAD-7: Generalized Anxiety Disorder-7
GBA: Gut Brain Axis
GI: Gastrointestinal
MDD: Major Depressive Disorder
MI: Motivational Interviewing
O3FA: Omega-3 Fatty Acid
PDA: Perinatal Depression and Anxiety
PPAQ: Physical Activity Questionnaire
RCT: Randomized Control Trial
REDCap: Research Electronic Data Capture
SCFA: Short Chain Fatty Acid
SISQS: Single Item Sleep Quality Scale

## Author’s Contributions

Conceptualization: NE, SG, KC

Methodology: NE, SG, KC

Writing (original): FG

Writing (review and editing): FG, NE, KC

Funding acquisition: NE, SG, KC

All authors read and approved the final transcript.

### Funding

This study was funded by the Lotte & John Hecht Memorial Foundation.

### Availability of Data and Materials

The datasets used and/or analyzed during the current study are available from the corresponding author on reasonable request.

