## Supplementary material for "Targeting Maternal Gut Microbiome to Improve Mental Health Outcomes-A Pilot Feasibility Trial": Randomization Flow Chart

### Randomization/Assignment or Exclusion Flowchart

#### [Based on fish oil & probiotic use at screening]

**Flowchart 1: P**atients taking BOTH or NEITHER Supplement

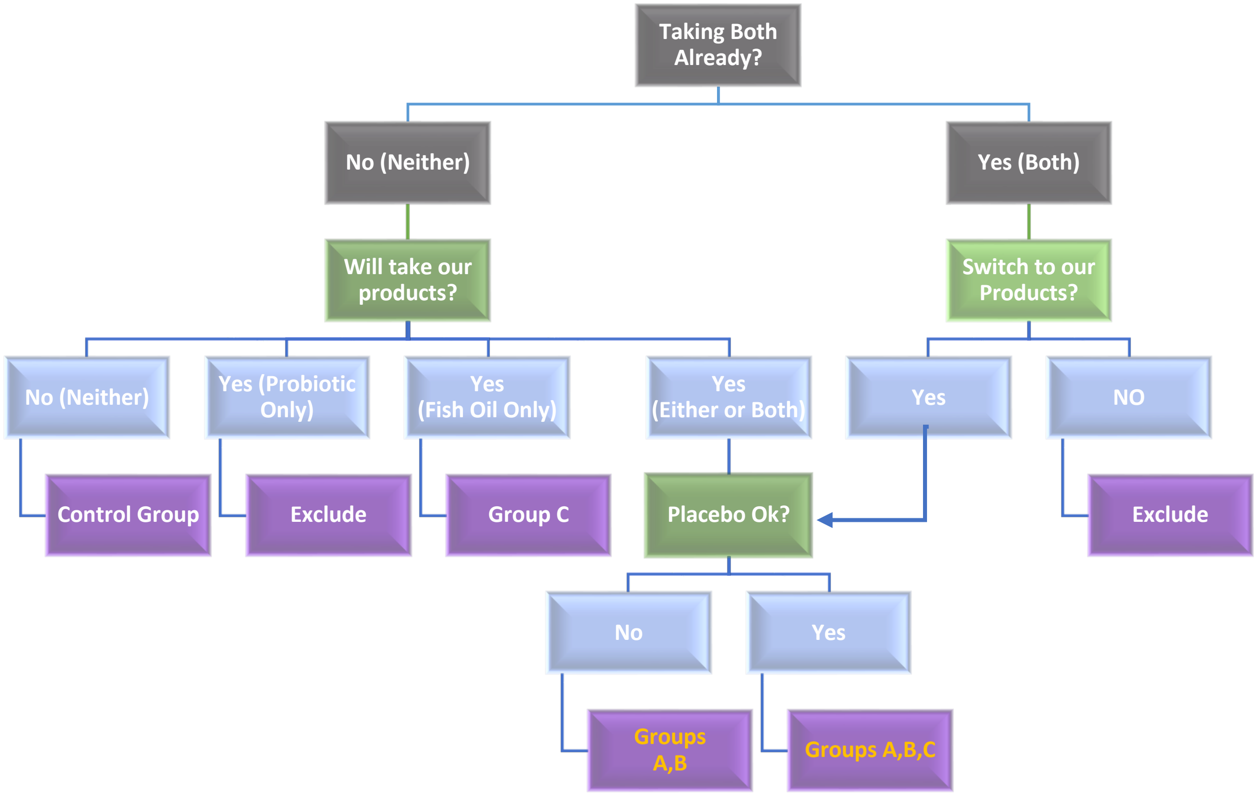

**Flow Chart 2:** Patients Preferring Fish Oil Only

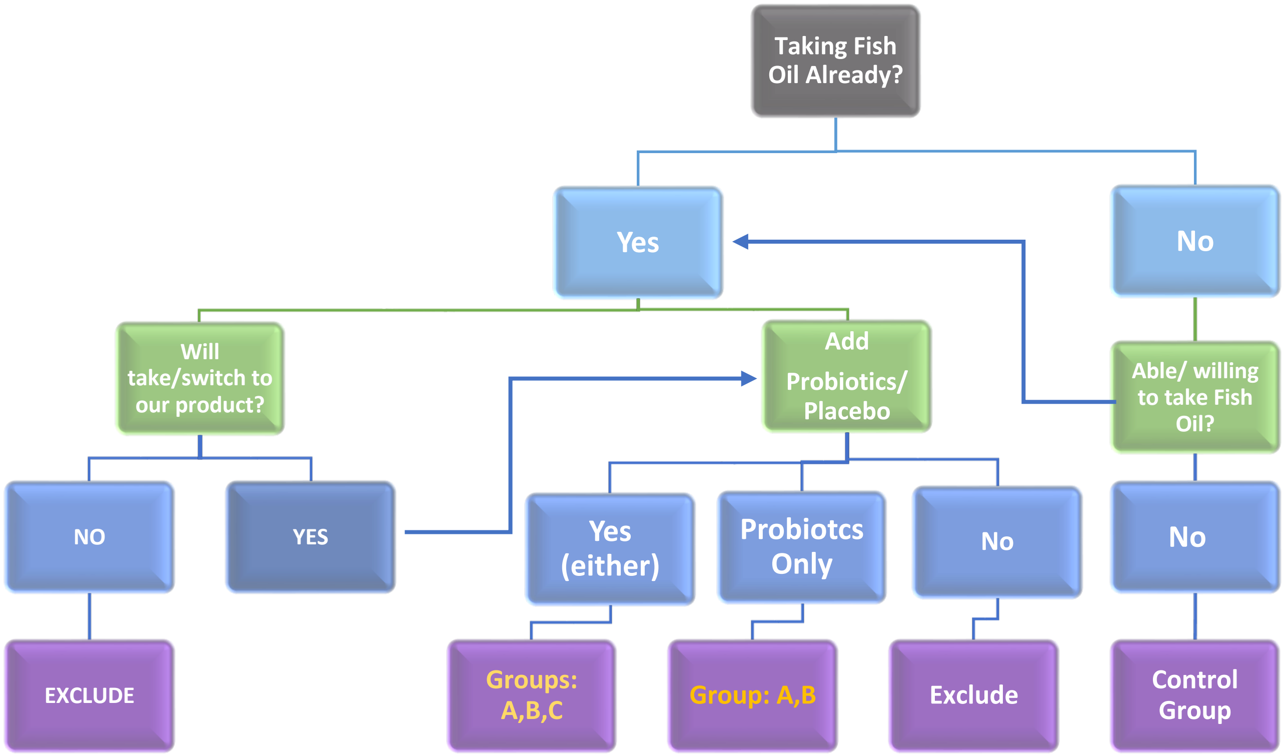

**Flowchart 3:** Patients Preferring Probiotics Only

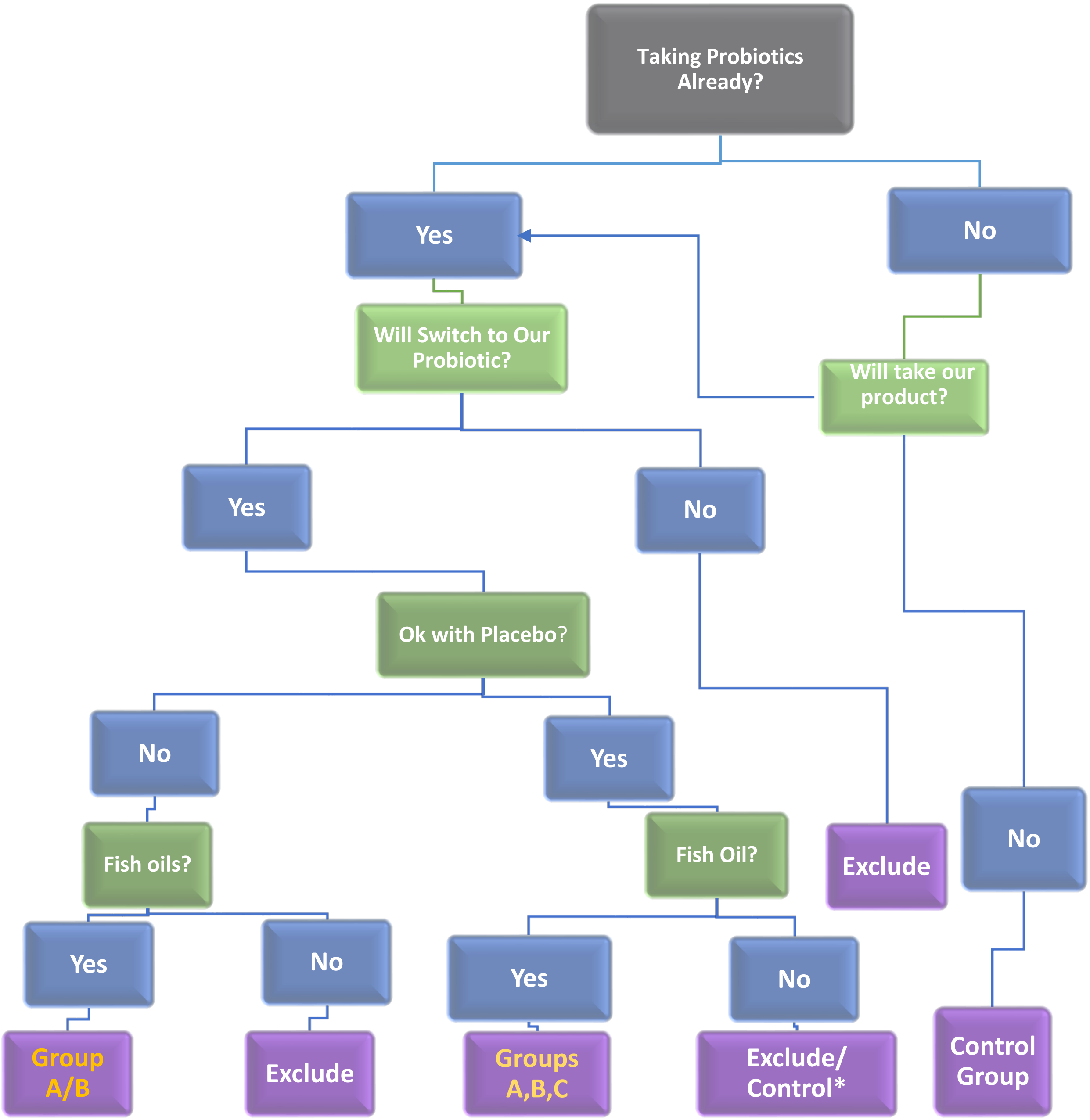

* For patients interested in placebo, but not fish oils, we will offer the option of being part of the active control arm.
To account for patient preference, full randomization to one of the three intervention groups is possible only if subjects have no objection to being in either of the groups.

- Subjects already on both or either of probiotics/Fish oils can only be randomized fully if they have no objection to modifying their supplements to include Fish oils and possibly being on placebo
- Subjects adamant about staying on Fish oils only, cannot be randomized and will be placed in Group C
- Subjects adamant about continuing probiotics but no fish oils, will be excluded.

Randomization status based on fish oil/probiotic responses on screening questionnaires.

| Fish Oil | Probiotics | Placebo | Group Assignment | Randomize |
|---|---|---|---|---|
| + | + | + | A,B,C | Yes |
| + | - | - | C | no |
| + | + | - | A,B | Yes |
| - | + | + | Control/Exclude | No |
| - | - | - | Control | no |
